# Live Birth Rates Among Women With Multiple Sclerosis in Sweden, 2004 to 2022: A Population-Based Registry Study

**DOI:** 10.64898/2026.10.21.26366100

**Authors:** Timothy P. Copeland, Kyla A. McKay, Katharina Fink

**Author notes:** **Corresponding author:** Timothy P. Copeland, Department of Clinical Neuroscience, Karolinska Institutet, Stockholm, Sweden.

## Abstract

**Background:** Women with multiple sclerosis (MS) have fewer children than other women, but whether this narrowed as pregnancy evidence became reassuring and treatment options expanded is uncertain.

**Objective:** To compare live birth trends from 2004 to 2022 between women with MS and all Swedish women.

**Methods:** Linking the Swedish MS, National Patient, and Medical Birth Registers with national statistics, we compared age-standardised live birth rates, counted expected births at national rates, and estimated adjusted annual live-birth risk ratios in a matched comparison sample.

**Results:** Women with MS (n=10,452; 76,989 woman-years) had 3,577 children, about 1,200 (25%) fewer than expected (observed-to-expected ratio 0.75, 95% CI 0.72 to 0.77). Standardised rate ratios were below 1 annually (median 0.77). Observed-to-expected ratios were 0.73 (0.68 to 0.79) in 2004 to 2008 and 0.70 (0.65 to 0.75) in 2019 to 2022 (population-by-period interaction p=0.31). Age- and region-adjusted risk ratios ranged from 0.75 to 0.86 across periods (heterogeneity p=0.020), closest to 1 in 2009 to 2013.

**Conclusion:** Women with MS had about one quarter fewer live births than other Swedish women, and the difference was similar in the first and last periods.

## Introduction

Women diagnosed with multiple sclerosis (MS) during their childbearing years often seek advice on how the disease may affect reproductive planning, and counselling has evolved as evidence on pregnancy in MS has accumulated. In 1998, Confavreux et al.^1^ reported a decline in relapse activity during pregnancy, followed by an increase in the first three postpartum months. A nationwide Danish cohort found no association between pregnancy and long-term disability accumulation^2^. Advances in disease management, particularly with disease-modifying therapies (DMTs) during planned conception windows, have integrated family planning into standard MS care^3^. Nevertheless, women with MS remain more likely to be childless; for example, an Italian multicentre study of women who had completed their reproductive years found that 22% with MS were childless, compared to 13% of controls^4^.

Simonsen et al.^5^ and Moberg et al.^6^ reported lower birth rates among women with MS in Norway and Denmark. However, whether this difference has narrowed as treatment and counselling practices have evolved remains uncertain. Sweden is a useful setting for this question, given rapid changes in treatment practices, including increased off-label rituximab use by the mid-2010s, and substantial regional variability in first-line use of highly effective DMTs^7^. While Fink et al.^8^ examined obstetric complications among Swedish women with MS, they did not address trends in live birth rates. We compared annual age-standardised live birth rates among women with MS and the general Swedish population from 2004 to 2022, expressed the difference in numbers of children, and examined in a matched comparison sample whether it changed over time after adjustment for measured characteristics.

## Materials and methods

### Study population and data sources

Women with MS were identified through the Swedish MS Registry (SMSreg; approximately 86% of prevalent cases in 2025)^9,10^ and the National Patient Register (NPR), from which we used ICD-10 inpatient diagnoses (from 1997) and outpatient diagnoses (from 2001). MS was defined by SMSreg registration or by two or more outpatient or one or more inpatient ICD-10 G35 codes in the NPR, and all women meeting this definition were included. The MS index date was the earliest of the SMSreg diagnosis date, registration date, or first qualifying G35 code in the NPR; in this prevalent cohort, it marks the first identification of MS in these sources rather than disease onset (Supplementary Appendix A1).

Follow-up spanned 2004 to 2022; the start year allowed for incomplete outpatient specialist reporting in the first years of nationwide collection. Statistics Sweden (SCB) linked the registers using personal identity numbers, assigned to all Swedish residents^11^, and the investigators received pseudonymised individual-level extracts without access to identifiable data.

Live births were captured using the Medical Birth Register (MBR; approximately 98% coverage of births)^12^. National live birth counts by maternal age^13^ and female population counts by age^14^ were obtained from the SCB statistical database, with maternal age defined in both populations as the age attained during the calendar year (Supplementary Appendix A2). For the matched-cohort analysis, education and family disposable income were sourced from the Longitudinal Integrated Database for Health Insurance and Labour Market Studies (LISA), residence from the Total Population Register, comorbidities from the NPR (Supplementary Table S1), and parity from the MBR.

### Population-rate analysis

Residence was determined using migration-register intervals. Eligible person-time began at the later of the MS index date, immigration, or 1 January 2004, and ended at the earlier of emigration, death, or 31 December 2022; women could re-enter after returning to Sweden. The numerator included all live-born children recorded in the MBR during eligible person-time, and age was restricted to 18 to 45 years.

### Matched-cohort analysis

The matched-cohort sample was drawn from an existing cohort that matched each woman with MS to up to 10 women with no record of MS by birth year, sex, and region; this cohort was used solely as a sampling frame without conditioning on the matched set (Supplementary Appendix A3). Women contributed one woman-year for each calendar year they were alive, residing in Sweden on 1 January, aged 18 to 45 in completed years on that date, and past their matched index date (residence exclusions in Supplementary Table S2). Any live birth recorded in the MBR during the year was counted as an event. Women who met the MS definition after their matched index date were classified as having MS from 1 January of the year after their MS index date. Some women sampled as cases in that cohort did not meet this study’s MS definition and were retained as comparison women (5.5% of comparison woman-years); excluding them changed the fully adjusted risk ratios by about 0.01 or less.

### Covariates

All covariates in the matched-cohort analysis were time-varying: seven comorbidities identified in the NPR (depression, anxiety, diabetes, hypertension, obesity, thyroid disease, and epilepsy), education (elementary, high school, or post-secondary) and family disposable income quintile from the preceding calendar year, parity (prior live-birth deliveries), region, and age on 1 January in six groups based on conventional five-year bands (18 to 19, 20 to 24, 25 to 29, 30 to 34, 35 to 39, and 40 to 45 years). Missing education, income, and region values were filled from the same woman’s other years, otherwise classified as unknown (Supplementary Appendix A4).

### Statistical analysis

For the population-rate analysis, annual live birth rates per 1,000 woman-years were calculated in the six age strata and directly standardised to the 2014 Swedish female population (Supplementary Tables S3 and S4), with gamma (Fay-Feuer) confidence intervals for standardised rates and the method of variance estimates recovery (MOVER) for their ratio. Expected live births were obtained by applying the national rate for each calendar year and age stratum to the person-time of women with MS. We calculated the ratio of observed to expected births, with exact Poisson confidence intervals, overall and for 2004 to 2008, 2009 to 2013, 2014 to 2018, and 2019 to 2022, and summed each period’s age-specific rates over ages 18 to 45, giving the number of children per woman those rates imply. To test whether trends differed between populations, we fitted a Poisson model to the age-stratum counts of both populations with a common age effect and an interaction between population and calendar time, specified once with year as a continuous term and once with categorical period. We described maternal age and disease duration by period for all deliveries, and the Expanded Disability Status Scale (EDSS) score before delivery and DMT exposure at conception for deliveries linked to SMSreg (Table 1; Supplementary Appendix A5).

**Table 1.** Characteristics of live-birth deliveries to women with MS, by calendar period.

| **No. (Column %), Mean±SD, or Median (Q1, Q3)** | **Total**<br>**N=3,524** | **2004-2008**<br>**N=715** | **2009-2013**<br>**N=972** | **2014-2018**<br>**N=1,039** | **2019-2022**<br>**N=798** |
| --- | --- | --- | --- | --- | --- |
| Mother's age at delivery (years) | 32 (29, 35) | 32 (29, 35) | 32 (29, 35) | 32 (29, 36) | 32 (30, 36) |
| MS duration (years) | 6±4 | 5±3 | 6±4 | 6±4 | 7±4 |
| MS duration group |  |  |  |  |  |
| <5 years | 1,677 (48) | 425 (59) | 446 (46) | 452 (44) | 354 (44) |
| 5-9 years | 1,279 (36) | 229 (32) | 385 (40) | 393 (38) | 272 (34) |
| 10+ years | 568 (16) | 61 (9) | 141 (15) | 194 (19) | 172 (22) |
| Recorded DMT episode within 30 days of estimated conception | 1,714 (52) | 146 (22) | 402 (45) | 597 (61) | 569 (76) |
| DMT Category |  |  |  |  |  |
| None | 1,555 (48) | 504 (78) | 492 (55) | 377 (39) | 182 (24) |
| Platform injectable | 693 (21) | 139 (21) | 308 (34) | 190 (20) | 56 (7) |
| Anti-CD20 | 543 (17) | 0 (0) | 8 (1) | 189 (19) | 346 (46) |
| Natalizumab | 359 (11) | 7 (1) | 86 (10) | 156 (16) | 110 (15) |
| Other | 119 (4) | 0 (0) | In Natalizumab | 62 (6) | 57 (8) |
| EDSS |  |  |  |  |  |
| 0-3.5 | 2,423 (74) | 338 (52) | 655 (73) | 779 (80) | 651 (87) |
| 4-5.5 | 91 (3) | 19 (3) | 31 (3) | 31 (3) | 10 (1) |
| 6+ | 40 (1) | 8 (1) | 12 (1) | 13 (1) | 7 (1) |
| Unknown | 715 (22) | 285 (44) | 196 (22) | 151 (16) | 83 (11) |
| Linked to MS Registry | 3,269 (93) | 650 (91) | 894 (92) | 974 (94) | 751 (94) |

Unit: delivery. EDSS and DMT are for deliveries linked to the MS Registry. EDSS is the most recent score before delivery, grouped at the scale's thresholds for walking impairment: 0-3.5, no limitation of walking range; 4-5.5, walking without aid but over a limited distance (from about 500 m at 4.0); 6+, a walking aid needed to walk about 100 m. MS duration: years from the MS index date to delivery. DMT: None, no recorded DMT episode within 30 days of estimated conception; Platform injectable, interferon beta or glatiramer acetate; Anti-CD20, rituximab, ocrelizumab, ofatumumab, or ublituximab; Other (oral, S1P-modulator, and other agents) is included in Natalizumab in 2009-2013 because of small counts. DMT, disease-modifying therapy; EDSS, Expanded Disability Status Scale; MS, multiple sclerosis; Q1 to Q3, first to third quartile; S1P, sphingosine-1-phosphate; SD, standard deviation.

For the matched-cohort analysis, we used modified Poisson regression with a log link and robust standard errors clustered on woman to estimate risk ratios (RRs) for a live birth within a calendar year among women with MS relative to the comparison sample^15^. The models have no person-time offset, so the RR compares annual probabilities of a live birth, not rates. All models included age group, region, calendar period, and an MS-by-period interaction, with a Wald test of equal period-specific RRs. To examine whether measured characteristics (Table 2) accounted for the difference, we added comorbidities, then education and income, and then parity in sequentially adjusted models. Analyses used Stata/SE 19.5 (StataCorp, College Station, TX); reporting follows the RECORD statement^16^.

**Table 2.**
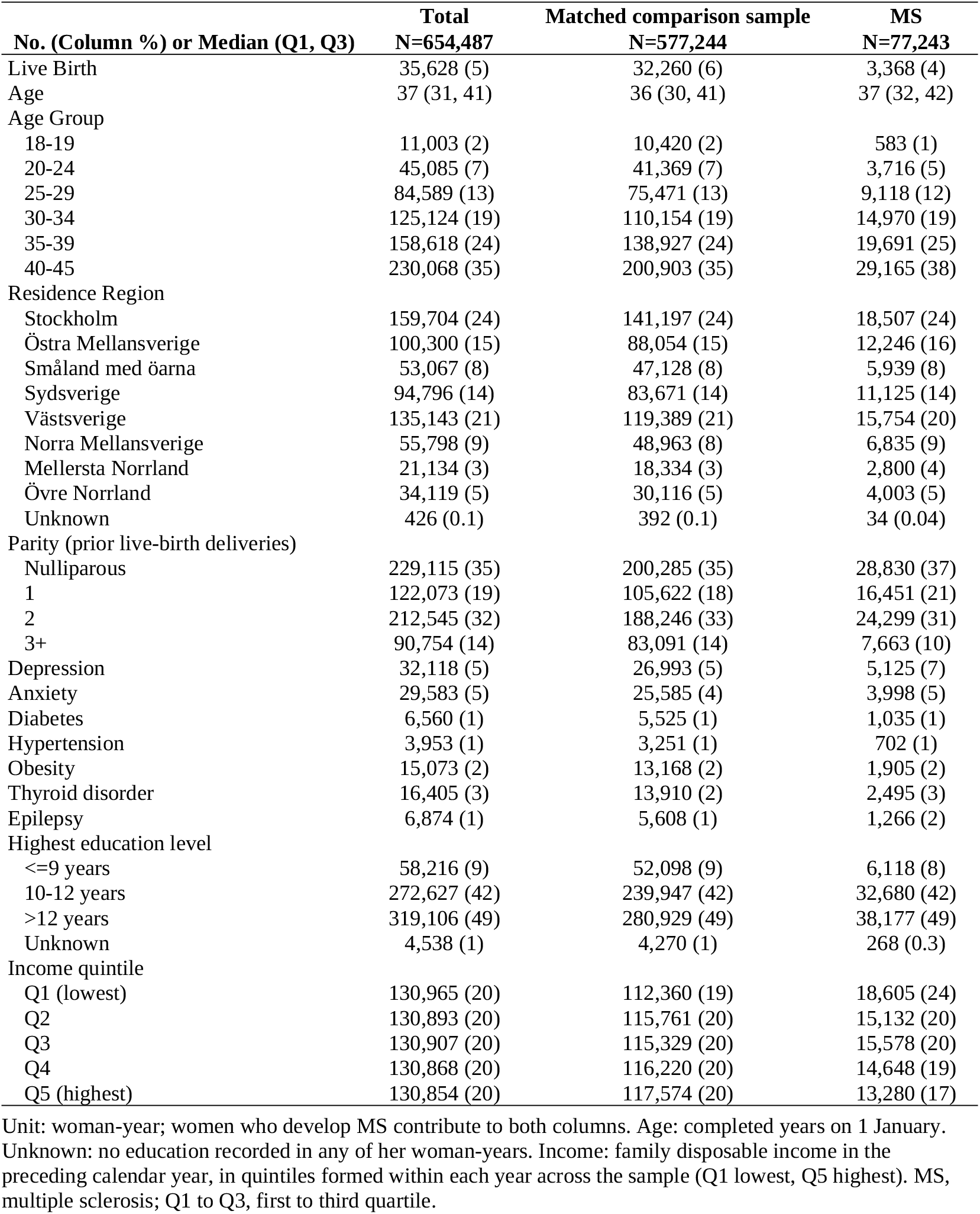
Woman-year characteristics by MS status in the matched comparison sample.

| **No. (Column %)** or **Median (Q1, Q3)** | **Total**<br>**N=654,487** | **Matched comparison sample**<br>**N=577,244** | **MS**<br>**N=77,243** |
| --- | --- | --- | --- |
| Live Birth | 35,628 (5) | 32,260 (6) | 3,368 (4) |
| Age | 37 (31, 41) | 36 (30, 41) | 37 (32, 42) |
| Age Group |  |  |  |
| 18-19 | 11,003 (2) | 10,420 (2) | 583 (1) |
| 20-24 | 45,085 (7) | 41,369 (7) | 3,716 (5) |
| 25-29 | 84,589 (13) | 75,471 (13) | 9,118 (12) |
| 30-34 | 125,124 (19) | 110,154 (19) | 14,970 (19) |
| 35-39 | 158,618 (24) | 138,927 (24) | 19,691 (25) |
| 40-45 | 230,068 (35) | 200,903 (35) | 29,165 (38) |
| Residence Region |  |  |  |
| Stockholm | 159,704 (24) | 141,197 (24) | 18,507 (24) |
| Östra Mellansverige | 100,300 (15) | 88,054 (15) | 12,246 (16) |
| Småland med öarna | 53,067 (8) | 47,128 (8) | 5,939 (8) |
| Sydsverige | 94,796 (14) | 83,671 (14) | 11,125 (14) |
| Västsverige | 135,143 (21) | 119,389 (21) | 15,754 (20) |
| Norra Mellansverige | 55,798 (9) | 48,963 (8) | 6,835 (9) |
| Mellersta Norrland | 21,134 (3) | 18,334 (3) | 2,800 (4) |
| Övre Norrland | 34,119 (5) | 30,116 (5) | 4,003 (5) |
| Unknown | 426 (0.1) | 392 (0.1) | 34 (0.04) |
| Parity (prior live-birth deliveries) |  |  |  |
| Nulliparous | 229,115 (35) | 200,285 (35) | 28,830 (37) |
| 1 | 122,073 (19) | 105,622 (18) | 16,451 (21) |
| 2 | 212,545 (32) | 188,246 (33) | 24,299 (31) |
| 3+ | 90,754 (14) | 83,091 (14) | 7,663 (10) |
| Depression | 32,118 (5) | 26,993 (5) | 5,125 (7) |
| Anxiety | 29,583 (5) | 25,585 (4) | 3,998 (5) |
| Diabetes | 6,560 (1) | 5,525 (1) | 1,035 (1) |
| Hypertension | 3,953 (1) | 3,251 (1) | 702 (1) |
| Obesity | 15,073 (2) | 13,168 (2) | 1,905 (2) |
| Thyroid disorder | 16,405 (3) | 13,910 (2) | 2,495 (3) |
| Epilepsy | 6,874 (1) | 5,608 (1) | 1,266 (2) |
| Highest education level |  |  |  |
| <=9 years | 58,216 (9) | 52,098 (9) | 6,118 (8) |
| 10-12 years | 272,627 (42) | 239,947 (42) | 32,680 (42) |
| >12 years | 319,106 (49) | 280,929 (49) | 38,177 (49) |
| Unknown | 4,538 (1) | 4,270 (1) | 268 (0.3) |
| Income quintile |  |  |  |
| Q1 (lowest) | 130,965 (20) | 112,360 (19) | 18,605 (24) |
| Q2 | 130,893 (20) | 115,761 (20) | 15,132 (20) |
| Q3 | 130,907 (20) | 115,329 (20) | 15,578 (20) |
| Q4 | 130,868 (20) | 116,220 (20) | 14,648 (19) |
| Q5 (highest) | 130,854 (20) | 117,574 (20) | 13,280 (17) |

Unit: woman-year; women who develop MS contribute to both columns. Age: completed years on 1 January. Unknown: no education recorded in any of her woman-years. Income: family disposable income in the preceding calendar year, in quintiles formed within each year across the sample (Q1 lowest, Q5 highest). MS, multiple sclerosis; Q1 to Q3, first to third quartile.

### Ethics

The Regional Ethical Review Board in Stockholm approved the study (2017/700-31/4), and the Swedish Ethical Review Authority approved an amendment (2021-02384). Individual informed consent was waived because the study used pseudonymised register data.

## Results

### Cohort and deliveries

The cohort included 20,910 women with MS, of whom 16,134 (77.2%) were registered in SMSreg, and 4,776 (22.8%) were identified through the NPR alone; 10,452 met the age and residence criteria in at least one year, contributing 76,989 resident woman-years. They had 3,524 live-birth deliveries comprising 3,577 live-born children, of which 100 (2.8%) were born in the calendar year of the MS index date. Median maternal age at delivery was 32 years (Q1 to Q3, 29 to 35). Between the first and last periods, mean MS duration at delivery increased from 5 (SD 3) to 7 (SD 4) years, and the proportion of deliveries among women with 10 or more years of disease increased from 9% to 22%. Of the 3,269 deliveries (93%) linked to SMSreg, EDSS was 0 to 3.5 for 74% and unrecorded for 22% (Table 1).

Among deliveries linked to SMSreg, the proportion with a recorded DMT episode within 30 days of estimated conception increased from 146 of 650 (22%) in 2004 to 2008 to 569 of 751 (76%) in 2019 to 2022 (Table 1); episodes without a recorded stop date were treated as continuing to the next treatment record or the end of 2022 (Supplementary Appendix A5), so later proportions are probably overstated. Platform injectables were the most common recorded exposure in each of the first three periods, peaking at 34% of linked deliveries in 2009 to 2013 and declining to 7% in 2019 to 2022, while anti-CD20 therapy increased from 1% in 2009 to 2013 to 46% in 2019 to 2022.

### Population-rate analysis

Among women with MS, the age-standardised live-birth rate was 42.4 per 1,000 woman-years in 2004 and 35.7 in 2022, ranging from 42 to 60 in the intervening years (highest in 2011 at 60.1, below 44 from 2019), with wide annual confidence intervals; general-population rates were greater throughout, increasing from 62.3 in 2004 to 70.0 in 2010 before declining to 54.3 in 2022 (Figure 1A; Supplementary Tables S3 and S4). The rate ratio was 0.68 in 2004 and 0.66 in 2022 (range 0.66 to 0.90); point estimates were below 1 each year, but the 95% CI included 1 in eight years (Figure 1B). General-population rates declined by an average 0.6% annually (incidence rate ratio [IRR] 0.994, 95% CI 0.991 to 0.997); the MS trend did not differ detectably (MS-by-year interaction IRR 1.000, 95% CI 0.992 to 1.009, p=0.97; categorical MS-by-period p=0.31; Supplementary Appendix A6).

**Figure 1.**
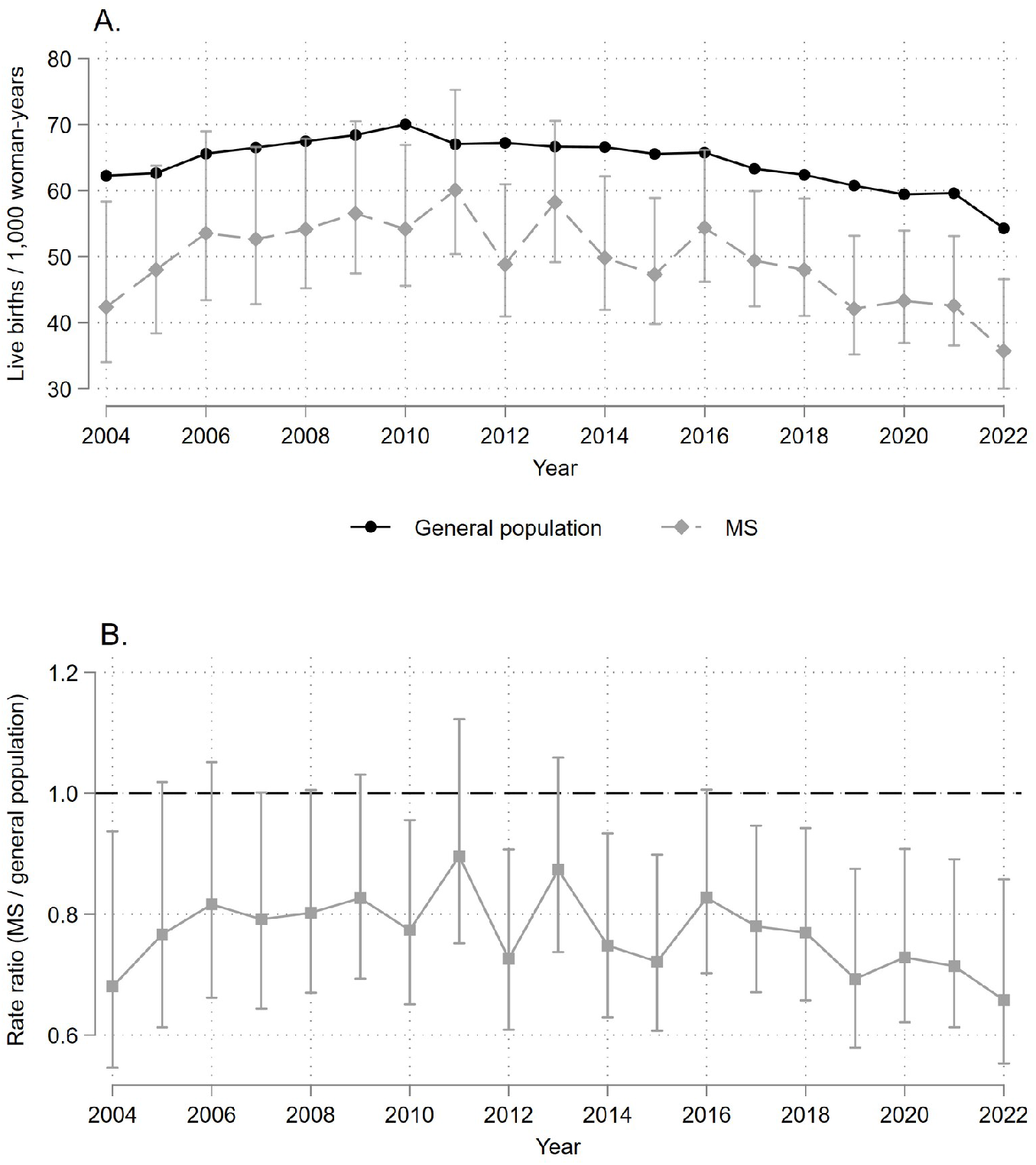
Age-standardised live birth rates and the MS-to-general-population rate ratio, Sweden, 2004 to 2022. Panel A plots annual live birth rates per 1,000 woman-years for the general population (solid line, circles) and for women with MS (dashed line, diamonds), with gamma (Fay-Feuer) 95% confidence intervals shown for the MS series. Panel B plots the ratio of the two standardised rates (solid grey line, squares) with 95% limits by the method of variance estimates recovery (MOVER), against a reference line at 1. Rates are directly standardised to the 2014 Swedish female population across six age bands, with age defined as calendar year minus year of birth (the age attained at year end) in both populations; annual values underlying both panels are in Supplementary Table S4.

Women with MS had 3,577 live-born children, against 4,801 expected at national age-specific rates, 1,224 (25%) fewer (observed-to-expected ratio 0.75, 95% CI 0.72 to 0.77; Table 3). The ratio was 0.73 (0.68 to 0.79) in 2004 to 2008, 0.79 (0.74 to 0.84) in 2009 to 2013, 0.76 (0.71 to 0.80) in 2014 to 2018, and 0.70 (0.65 to 0.75) in 2019 to 2022. The shortfall rose from about 50 children a year in 2004 to 2008 to about 90 a year in 2019 to 2022, mainly because annual person-time among women with MS grew by about half. The 2019 to 2022 age-specific rates sum to 1.16 children per woman over ages 18 to 45 among women with MS and 1.65 nationally, a summary of period rates, not completed family size.

**Table 3.** Live births among women with MS by calendar period: observed and expected numbers, children per woman, and annual risk ratios against the matched comparison sample.

|  | **2004-2008** | **2009-2013** | **2014-2018** | **2019-2022** | **2004-2022** |
| --- | --- | --- | --- | --- | --- |
| MS live-born children, observed | 727 | 986 | 1,056 | 808 | 3,577 |
| MS live-born children, expected at national rates | 994 | 1,251 | 1,397 | 1,159 | 4,801 |
| Fewer than expected, n (%) | 267 (27) | 265 (21) | 341 (24) | 351 (30) | 1,224 (25) |
| Observed/expected (95% CI) | 0.73 (0.68 to 0.79) | 0.79 (0.74 to 0.84) | 0.76 (0.71 to 0.80) | 0.70 (0.65 to 0.75) | 0.75 (0.72 to 0.77) |
| Children per woman aged 18-45, MS | 1.41 | 1.56 | 1.40 | 1.16 | 1.38 |
| Children per woman aged 18-45, national | 1.82 | 1.91 | 1.82 | 1.65 | 1.80 |
| Risk ratio (95% CI), base | 0.76 (0.70 to 0.82) | 0.86 (0.81 to 0.92) | 0.81 (0.76 to 0.86) | 0.75 (0.70 to 0.80) |  |
| Risk ratio (95% CI), fully adjusted | 0.73 (0.68 to 0.79) | 0.82 (0.77 to 0.87) | 0.79 (0.74 to 0.84) | 0.74 (0.69 to 0.80) |  |

Expected: national age- and year-specific rates applied to MS woman-years; fewer than expected: expected minus observed, with its percentage of expected; observed/expected with exact Poisson 95% CI. Children per woman aged 18-45: the period's age-specific rates summed over ages 18 to 45 (Supplementary Appendix A6). Risk ratio: annual probability of a live birth vs the matched comparison sample; the base model adjusts for age group and region, and the fully adjusted model adds comorbidities, education, income, and parity. p for equal risk ratios across periods: 0.020 (base), 0.048 (fully adjusted). CI, confidence interval; MS, multiple sclerosis.

### Matched-cohort analysis

The matched-cohort sample included 85,216 women and 654,487 woman-years (77,243 MS and 577,244 comparison), with a live birth in 4% of MS and 6% of comparison woman-years. Women with MS were older (median 37 vs 36 years) and more often nulliparous (37% vs 35%), with greater prevalences of depression (7% vs 5%), epilepsy (2% vs 1%), and thyroid disease (3.2% vs 2.4%); they were also more often in the lowest income quintile (24% vs 19%) (Table 2).

Women with MS had a lower annual probability of a live birth in every period and in every model (Table 3; Supplementary Table S5). Adjustment for comorbidities, education, and income changed the risk ratios by at most 0.02, and adjustment for parity moved them 0.03 to 0.05 further from 1. Across the four periods, RRs were 0.76, 0.86, 0.81, and 0.75 in the base model and 0.73, 0.82, 0.79, and 0.74 when fully adjusted (95% CIs in Table 3). A joint test of equal RRs across periods gave p=0.020 in the base model and 0.048 when fully adjusted; the RRs were closest to 1 in 2009 to 2013 and similar in the first and last periods. In the comparison sample, the adjusted live-birth probability was 7% lower in 2019 to 2022 than in 2004 to 2008 (RR 0.93, 0.90 to 0.95; Supplementary Table S6).

## Discussion

Women with MS in Sweden had live birth rates about one quarter lower than the general population from 2004 to 2022 (observed-to-expected ratio 0.75), approximately 1,200 fewer children than expected. National rates rose until 2010 then declined, consistent with the broader decline in Nordic birth rates^17^; rates among women with MS varied more from year to year and were lowest in 2019 to 2022. The population interaction tests did not detect a difference in trends (period p=0.31), although standard errors clustered on calendar year gave p=0.032 (Supplementary Appendix A6). In the matched sample, risk ratios differed across periods (p=0.020) but varied rather than narrowed.

Over the study period, evidence accumulated that pregnancy does not worsen long-term MS course^2^, and more DMT options compatible with pregnancy planning became available, including planned pauses of anti-CD20 therapy around conception^18^. Recorded DMT exposure near conception rose from 22% to 76% of SMSreg-linked deliveries, although the later figure is probably overstated. These developments may have been expected to narrow the gap, but it was similar in the first and last periods. Part of the difference may reflect lower fertility that begins before diagnosis and persists after it, given pregnancy-related health care use is already lower during the MS prodrome^19^ and birth rates in Norway were reduced from the prodrome onwards^5^.

The lower annual rates among women with MS may reflect voluntary childlessness, reduced fecundity, pregnancy loss or termination, or deferral of pregnancy for MS treatment; because only live births are recorded, lower conception cannot be separated from pregnancy loss or termination. Among Danish women receiving assisted reproduction, the chance of a live birth per embryo transfer was similar with and without MS (adjusted odds ratio 0.91, 95% CI 0.81 to 1.02)^20^, but because these women had already sought fertility treatment, this does not show whether women with MS conceive less readily. Women with MS who have children also face postpartum demands, including relapse risk in the first months after delivery^1,21^ and an increased incidence of peripartum depression and other mental illness^22,23^. Counselling on reproduction appears incomplete. In a 2016 to 2017 volunteer online survey of Danish women with MS, 42% did not know whether any DMT could affect the fetus, and 41% named their neurologist as a source of information on fetal risk^24^.

Adjustment for parity moved the risk ratios further from 1 because women with MS were more often in the group with one prior delivery (21% vs 18% of woman-years), which had the greatest annual probability of a live birth, and less often in the group with two or more (41% vs 47%), which had the lowest. Because parity counts earlier births, some after diagnosis, it partly reflects the difference under study, so the fully adjusted RRs compare women with equal numbers of earlier births rather than giving a more complete estimate of the overall difference.

Consistently lower rates have also been reported in Norway^5^, Denmark^6^, and the Campania region of Italy, where the fertility rate among women with MS was less than half the regional rate in 2018 to 2020^25^. In United States claims data, by contrast, the adjusted proportion of women with MS who were pregnant in a year rose from 7.9% in 2006 to 9.5% in 2014 while it fell from 8.8% to 7.8% among women without MS^26^, a convergence not present in this study.

This study covered all registry-identified women with MS over 19 years and compared them with both the national female population and a matched sample with individual-level covariates. For 93% of deliveries, linkage to SMSreg provided DMT information at conception, and 78% of these had an EDSS score before delivery. The findings describe a prevalent, registry-identified population with tax-funded health care and may not apply where access to MS or fertility care differs.

For women identified only through outpatient care, the index date is the first of the two qualifying visits, so some person-time precedes diagnosis confirmation: 235 of 76,989 woman-years (0.3%) and 17 live-born children, too few to materially change the population rates. The index date can also precede SMSreg registration, so early post-index person-time is included only for women who later registered or met the NPR definition. Country of birth was not accounted for in either comparison; if foreign-born women, a growing share of Swedish women of reproductive age, have both higher fertility and a lower prevalence of diagnosed MS, national rates would slightly overstate the births expected for women with MS. Revisions of the McDonald criteria from 2001 to 2017 moved diagnosis earlier and towards milder disease, a change in case mix that could mask or mimic a change in the difference over time.

Women with MS in Sweden had about one quarter fewer live births than other women, approximately 65 fewer children a year from 2004 to 2022, while evidence on pregnancy in MS became reassuring and therapies compatible with pregnancy planning expanded. The difference was similar in the earliest and latest periods and persisted after adjustment for comorbidity and socioeconomic position. Registers cannot establish whether this reflects informed choice or constraints such as reduced fecundity or incomplete counselling, which would call for different policy responses. Family planning could be discussed routinely in MS care, reviewed annually and considered when a DMT is chosen^3^, with access to maternity and fertility services; women with MS need not be advised against assisted reproduction^3^, and live birth after embryo transfer appears about as likely as in other women^20^. This may not close the gap but may help ensure informed decisions. Swedish registers allow the difference to be monitored as care changes.

## Supporting information

All supplementary material including RECORD checklist

## Data Availability

Individual-level data from the Swedish registers cannot be shared publicly under Swedish law. Researchers may apply for access through the Swedish National Board of Health and Welfare (Socialstyrelsen), Statistics Sweden (SCB), and the Swedish MS Registry, subject to ethical approval. The national birth and population counts are publicly available from the Statistics Sweden statistical database. Analysis code is available from the corresponding author on reasonable request. No study protocol was registered or published.

## Acknowledgements

We thank the women with MS and the clinicians who contribute data to the Swedish MS Registry. Claude (Anthropic) and GPT (OpenAI) were used to review the Stata code and to edit the manuscript text; TPC verified all changes.

## Statements and Declarations

### Ethical considerations

The study was approved by the Regional Ethical Review Board in Stockholm (2017/700-31/4), with an amendment approved by the Swedish Ethical Review Authority (2021-02384).

### Consent to participate

The requirement for individual informed consent was waived by the ethical review board because the study used pseudonymised register data.

### Consent for publication

Not applicable.

