## Supplementary material for "Live Birth Rates Among Women With Multiple Sclerosis in Sweden, 2004 to 2022: A Population-Based Registry Study": All supplementary material including RECORD checklist

**Abbreviations:** CI, confidence interval; DMT, disease-modifying therapy; EDSS, Expanded Disability Status Scale; ICD-10, International Classification of Diseases, 10th Revision; MBR, Medical Birth Register; MOVER, method of variance estimates recovery; MS, multiple sclerosis; NPR, National Patient Register; NUTS-2, second level of the Nomenclature of Territorial Units for Statistics; RR, risk ratio; SCB, Statistics Sweden; SES, socioeconomic status (education and income); SMSreg, Swedish MS Registry.

**Supplementary Appendix: Additional methods**

**A1. MS identification and index date.** The MS index date was the earliest of the SMSreg diagnosis date, the SMSreg registration date, the first inpatient G35 code in the NPR, and, for women with two or more outpatient G35 visits, the first of those visits; a single outpatient G35 visit did not contribute to the index date, even for women who met the MS definition through SMSreg or an inpatient code. When both SMSreg dates were missing, we used the SMSreg consent date. ICD-10 G35 codes in the NPR are available from 1997 for inpatient and 2001 for outpatient records, so for women identified only through the NPR, the index date, and therefore disease duration, cannot precede those years. Because the cohort is prevalent, the index date marks the first record of MS in these sources rather than disease onset, and women identified before 2004 contributed person-time from 1 January 2004. All 20,910 women with MS linked to the SCB demographic file with a recorded birth date. Of the 16,134 women in SMSreg, 212 (1.3%) were recorded as not fulfilling the diagnostic criteria and 155 (1.0%) had no entry for this field; 139 and 123 of these, respectively, also met the NPR definition. The remaining 105 (0.5% of the cohort) were included on SMSreg registration alone, because the study definition does not use the criteria field. We found no published validation study of this case definition in SMSreg or the NPR, and none was done for this study; the SMSreg diagnostic-criteria field is the only internal check of case status.

**A2. Population rates.** As in SCB age-specific fertility rates, maternal age was the age attained during the calendar year (calendar year minus year of birth), and the national denominator for each year and age was the mean number of women of that birth year on 31 December of the preceding and current years. The MS numerator counted every live-born child recorded in the MBR during eligible person-time, so a twin delivery counted twice. The national rates include women with MS, but at a prevalence of 0.2% to 0.3% their effect on those rates is minimal. Supplementary Table S3 combines ages 18 to 19 and 20 to 24 by calendar period; the annual standardised rates in Figure 1 and Supplementary Table S4 use all six age groups.

**A3. Matched comparison sample.** The matching frame matched each person with MS to up to 10 people with no record of MS on birth year, sex, and region, and assigned controls the index date of their case. Restricted to women, it held 31,355 women sampled as cases and 153,397 controls (a mean of 4.9 per case), each appearing once. We used the frame only as a source of comparison women and did not condition on the matched set. A woman contributed a woman-year if, on 1 January, she was alive, registered as resident in Sweden, aged 18 to 45 in completed years, and past her matched index date; gaps and re-entry were allowed. Supplementary Table S2 gives the proportion of otherwise eligible woman-years excluded by this 1 January residence check. The population-rate analysis instead followed residence continuously through migration-register intervals, with no 1 January check and so no such exclusions. All 184,752 frame women linked to the SCB demographic file with a recorded birth date and index date. Of their 3,510,288 calendar years over 2004 to 2022, 2,837,142 were excluded because the woman was outside ages 18 to 45 on 1 January or the year did not follow her index year, 2,811 because she had died before 1 January, and 15,848 by the residence check, leaving 654,487 woman-years from 85,216 women; the other 99,536 frame women had no eligible woman-year. The frame did not include 102 women in the MS cohort, who were therefore absent from the matched-cohort analysis only. Follow-up was not censored within the year, so women who died, emigrated, or turned 46 after 1 January remained in that year’s risk set. Women who met the study MS definition contributed comparison woman-years up to and including their index year and MS woman-years from 1 January of the following year. Fewer than five frame controls ever met the definition, so these were almost all women sampled as cases whose study index year followed their frame index year. Of the comparison woman-years, 5.5% came from women sampled as cases who did not meet the study definition and 0.4% from women with MS before reclassification. Their crude annual live-birth proportions (5.2% and 6.4%) were close to that of frame controls (5.6%), and excluding the former changed the fully adjusted RRs by about 0.01 or less.

**A4. Covariates.** Each comorbidity required two qualifying ICD-10 codes on separate dates in inpatient or outpatient records and was present from 1 January of the year after the second code (Supplementary Table S1). Because ICD-10 records begin in 1997 (inpatient) and 2001 (outpatient), the look-back lengthens over calendar time, and recorded comorbidity prevalence rises in both groups for that reason alone. Education in the preceding calendar year had three levels: elementary (at most 9 years), high school (10 to 12 years), and post-secondary (more than 12 years). Family disposable income in the preceding year was grouped into quintiles of the pooled sample, formed within each calendar year so that income growth over the study period did not move women between groups. Region was the county of residence in the Total Population Register for the same calendar year, grouped into the eight NUTS-2 regions. Parity counted prior live-birth deliveries in the MBR, with a twin delivery counted once. Missing education, income, and region were filled from the same woman's other years, carrying the next observation backward and then the last observation forward for education and region, and in the reverse order for income. After filling, income was complete, and education (0.7% of woman-years) and region (fewer than 0.1%) were classified as unknown for women with no value in any year. Age in the matched-cohort analysis was completed years on 1 January, so its age groups are about one year younger than those of the population-rate analysis (calendar year minus year of birth).

**A5. EDSS and DMT at conception.** For deliveries linked to SMSreg, EDSS was the most recent score before delivery (unknown if none was recorded), grouped as 0-3.5, 4.0-5.5 and 6.0 or higher at the scale's thresholds for limited walking range (4.0) and need for a walking aid (6.0), and DMT exposure at conception came from SMSreg treatment episodes, excluding symptomatic treatment, records of no treatment or of other non-disease-modifying treatment, and study drugs. Estimated conception was the delivery date minus gestational age plus 14 days. A delivery was exposed if an episode was ongoing at estimated conception or started or ended within 30 days of it, with DMT class taken from the episode closest to conception or, if more than one was ongoing, the most recently started. Episodes without a stop date, which the registry flags as ongoing, were assumed to continue until the next treatment, no-treatment, or study-drug record or the end of 2022, except for treatments given once or in short courses (alemtuzumab, cladribine, mitoxantrone, cyclophosphamide, and autologous haematopoietic stem cell transplantation), which counted only on the treatment date. In Table 1 the unit is the delivery (a twin delivery counts once), and the Total column keeps the 2009 to 2013 deliveries with Other DMT within natalizumab.

**A6. Statistical details.** Expected births were the sum, over calendar year and age strata, of the national rate multiplied by the woman-years of women with MS (indirect standardisation). Observed-to-expected ratios have exact Poisson confidence intervals that treat the expected count as fixed. The gamma and MOVER intervals treat live-born children as independent events and the two standardised rates as independent, so they ignore twin deliveries, repeat births to one woman, and the inclusion of women with MS in the national rate. In Table 3, "fewer than expected" is expected minus observed births, with its percentage of expected; the observed and expected counts and children per woman come from the population-rate analysis, which counts person-time, and the risk ratios from the matched-cohort analysis, which counts woman-years (Supplementary Appendix A3). Children per woman is the sum over the six age groups of the annual standardisation (18 to 19, 20 to 24, 25 to 29, 30 to 34, 35 to 39, and 40 to 45 years) of each group's live-birth rate for the period multiplied by the number of years the group spans (2, 5, 5, 5, 5, and 6). It cannot be reproduced exactly from Supplementary Table S3, which combines ages 18 to 24, and it summarises the period's rates, not the completed family size of women with MS. The national figure differs from the official total fertility rate, which covers ages 15 to 49. The population trend models used a log person-time offset and robust standard errors and treated the 228 age-year-population cells as independent. The MS-by-year term tests only a linear difference in trend and cannot detect a rise and fall; the categorical MS-by-period term can. With standard errors clustered on calendar year, the MS-by-period test gave p=0.032, though clustering can understate uncertainty with only 19 clusters. p values are not adjusted for the several trend and interaction tests; we interpret them alongside the confidence intervals rather than against a fixed threshold. The base and fully adjusted risk ratios in Table 3 are the first and last of the four sequential models in Supplementary Table S5. Supplementary Table S6 gives every estimate from the fully adjusted model; its calendar-period rows are risk ratios within the comparison sample, holding the other covariates fixed.

### Supplementary Tables

#### Table S1. ICD-10 codes used to ascertain comorbidities from the National Patient Register.

| Comorbidity | ICD-10 codes |
|---|---|
| Depression | F32, F33 |
| Anxiety | F40, F41 |
| Diabetes | E10–E14 |
| Hypertension | I10–I13, I15 |
| Obesity | E66 |
| Thyroid disorder | E00–E07 |
| Epilepsy | G40 |

Two codes on separate dates, inpatient or outpatient; present from 1 January of the year after the second code.

### Table S2. Woman-years excluded by the annual 1 January residence check in the matched-cohort analysis.

| Year | Excluded / eligible (%) |
|---|---|
| 2004 | 192 / 20,231 (0.9) |
| 2005 | 259 / 22,554 (1.1) |
| 2006 | 332 / 24,861 (1.3) |
| 2007 | 421 / 26,610 (1.6) |
| 2008 | 521 / 28,870 (1.8) |
| 2009 | 581 / 30,655 (1.9) |
| 2010 | 627 / 32,194 (1.9) |
| 2011 | 690 / 33,361 (2.1) |
| 2012 | 749 / 34,535 (2.2) |
| 2013 | 844 / 35,823 (2.4) |
| 2014 | 949 / 37,485 (2.5) |
| 2015 | 1,025 / 38,677 (2.7) |
| 2016 | 1,118 / 40,050 (2.8) |
| 2017 | 1,168 / 41,710 (2.8) |
| 2018 | 1,174 / 42,801 (2.7) |
| 2019 | 1,217 / 43,520 (2.8) |
| 2020 | 1,290 / 44,566 (2.9) |
| 2021 | 1,312 / 45,412 (2.9) |
| 2022 | 1,379 / 46,420 (3.0) |

Excluded / eligible (%): woman-years excluded because the woman was not registered as resident in Sweden on 1 January, over the woman-years that met all other criteria (alive, aged 18 to 45, and past the matched index date), with the percentage excluded in parentheses.

**Table S3. Age-specific live birth rates per 1,000 woman-years by calendar period.**

| Calendar period | Age group (years) | MS woman-years | MS live-born children | MS rate per 1,000 | General-population woman-years | General-population live births | General-population rate per 1,000 | 2014 standard population |
|---|---|---|---|---|---|---|---|---|
| 2004-2008 | 18-24 | 832.7 | 29 | 34.8 | 1,898,919 | 61,659 | 32.5 | 434,874 |
| | 25-29 | 1,802.3 | 157 | 87.1 | 1,340,166 | 141,943 | 105.9 | 320,517 |
| | 30-34 | 3,113.6 | 324 | 104.1 | 1,468,293 | 194,410 | 132.4 | 297,018 |
| | 35-39 | 4,250.5 | 180 | 42.3 | 1,553,991 | 101,979 | 65.6 | 296,259 |
| | 40-45 | 6,386.0 | 37 | 5.8 | 1,883,521 | 23,176 | 12.3 | 382,478 |
| 2009-2013 | 18-24 | 1,026.1 | 36 | 35.1 | 2,180,924 | 71,181 | 32.6 | 434,874 |
| | 25-29 | 2,164.5 | 201 | 92.9 | 1,444,720 | 155,332 | 107.5 | 320,517 |
| | 30-34 | 3,616.4 | 402 | 111.2 | 1,429,036 | 195,557 | 136.8 | 297,018 |
| | 35-39 | 5,002.9 | 283 | 56.6 | 1,528,880 | 114,392 | 74.8 | 296,259 |
| | 40-45 | 7,654.7 | 64 | 8.4 | 1,926,369 | 27,628 | 14.3 | 382,478 |
| 2014-2018 | 18-24 | 1,151.2 | 28 | 24.3 | 2,039,474 | 59,832 | 29.3 | 434,874 |
| | 25-29 | 2,614.7 | 229 | 87.6 | 1,701,827 | 174,975 | 102.8 | 320,517 |
| | 30-34 | 4,229.0 | 424 | 100.3 | 1,554,544 | 200,794 | 129.2 | 297,018 |
| | 35-39 | 5,590.2 | 290 | 51.9 | 1,506,279 | 111,738 | 74.2 | 296,259 |
| | 40-45 | 8,431.5 | 85 | 10.1 | 1,902,465 | 29,211 | 15.4 | 382,478 |
| 2019-2022 | 18-24 | 910.1 | 14 | 15.4 | 1,525,825 | 33,123 | 21.7 | 434,874 |
| | 25-29 | 2,250.7 | 153 | 68.0 | 1,357,897 | 122,288 | 90.1 | 320,517 |
| | 30-34 | 3,952.0 | 349 | 88.3 | 1,453,438 | 174,135 | 119.8 | 297,018 |
| | 35-39 | 4,905.6 | 233 | 47.5 | 1,297,823 | 91,681 | 70.6 | 296,259 |
| | 40-45 | 7,104.5 | 59 | 8.3 | 1,500,275 | 23,916 | 15.9 | 382,478 |

Age: calendar year minus year of birth. Ages 18-24 are combined so that no MS cell has fewer than 5 mothers.

**Table S4. Annual crude and age-standardised live birth rates per 1,000 woman-years.**

| Year | MS woman-years | MS live-born children | Crude MS rate | Standardised MS rate (95% CI) | General-population woman-years | General-population live births | Crude general-population rate | Standardised general-population rate (95% CI) | Rate ratio (95% CI) |
|---|---|---|---|---|---|---|---|---|---|
| 2004 | 2,976 | 124 | 41.7 | 42.4 (34.0 to 58.3) | 1,604,794 | 100,612 | 62.7 | 62.3 (61.9 to 62.7) | 0.68 (0.55 to 0.94) |
| 2005 | 3,145 | 130 | 41.3 | 48.0 (38.4 to 63.8) | 1,611,426 | 101,011 | 62.7 | 62.6 (62.3 to 63.0) | 0.77 (0.61 to 1.02) |
| 2006 | 3,286 | 144 | 43.8 | 53.6 (43.4 to 69.0) | 1,624,217 | 105,556 | 65.0 | 65.6 (65.2 to 66.0) | 0.82 (0.66 to 1.05) |
| 2007 | 3,421 | 152 | 44.4 | 52.7 (42.8 to 66.6) | 1,641,750 | 107,037 | 65.2 | 66.5 (66.1 to 66.9) | 0.79 (0.64 to 1.00) |
| 2008 | 3,557 | 177 | 49.8 | 54.2 (45.2 to 67.9) | 1,662,704 | 108,951 | 65.5 | 67.5 (67.1 to 67.9) | 0.80 (0.67 to 1.01) |
| 2009 | 3,730 | 184 | 49.3 | 56.6 (47.4 to 70.5) | 1,682,768 | 111,410 | 66.2 | 68.4 (68.0 to 68.8) | 0.83 (0.69 to 1.03) |
| 2010 | 3,803 | 197 | 51.8 | 54.2 (45.6 to 66.9) | 1,696,211 | 115,247 | 67.9 | 70.0 (69.6 to 70.4) | 0.77 (0.65 to 0.96) |
| 2011 | 3,879 | 202 | 52.1 | 60.1 (50.4 to 75.3) | 1,704,617 | 111,415 | 65.4 | 67.1 (66.7 to 67.4) | 0.90 (0.75 to 1.12) |
| 2012 | 3,963 | 183 | 46.2 | 48.8 (40.9 to 61.0) | 1,710,963 | 112,801 | 65.9 | 67.2 (66.8 to 67.6) | 0.73 (0.61 to 0.91) |
| 2013 | 4,090 | 220 | 53.8 | 58.2 (49.2 to 70.6) | 1,715,370 | 113,217 | 66.0 | 66.7 (66.3 to 67.1) | 0.87 (0.74 to 1.06) |
| 2014 | 4,167 | 195 | 46.8 | 49.8 (41.9 to 62.2) | 1,721,176 | 114,543 | 66.5 | 66.6 (66.2 to 67.0) | 0.75 (0.63 to 0.93) |
| 2015 | 4,279 | 194 | 45.3 | 47.3 (39.8 to 58.9) | 1,728,534 | 114,505 | 66.2 | 65.6 (65.2 to 65.9) | 0.72 (0.61 to 0.90) |
| 2016 | 4,407 | 225 | 51.1 | 54.4 (46.2 to 66.2) | 1,739,112 | 117,036 | 67.3 | 65.8 (65.4 to 66.1) | 0.83 (0.70 to 1.01) |
| 2017 | 4,547 | 227 | 49.9 | 49.4 (42.5 to 59.9) | 1,752,153 | 115,008 | 65.6 | 63.3 (63.0 to 63.7) | 0.78 (0.67 to 0.95) |
| 2018 | 4,618 | 215 | 46.6 | 48.0 (41.0 to 58.8) | 1,763,614 | 115,458 | 65.5 | 62.4 (62.0 to 62.8) | 0.77 (0.66 to 0.94) |
| 2019 | 4,672 | 188 | 40.2 | 42.1 (35.2 to 53.2) | 1,773,065 | 114,126 | 64.4 | 60.8 (60.4 to 61.1) | 0.69 (0.58 to 0.87) |
| 2020 | 4,740 | 215 | 45.4 | 43.3 (36.9 to 53.9) | 1,778,018 | 112,695 | 63.4 | 59.4 (59.1 to 59.8) | 0.73 (0.62 to 0.91) |
| 2021 | 4,837 | 223 | 46.1 | 42.6 (36.6 to 53.1) | 1,785,140 | 113,971 | 63.8 | 59.6 (59.3 to 60.0) | 0.71 (0.61 to 0.89) |
| 2022 | 4,874 | 182 | 37.3 | 35.7 (30.0 to 46.6) | 1,799,035 | 104,351 | 58.0 | 54.3 (54.0 to 54.6) | 0.66 (0.55 to 0.86) |

Age: calendar year minus year of birth. Crude: live-born children per 1,000 woman-years without age standardisation. Standardised to the 2014 Swedish female population; gamma (Fay-Feuer) 95% CIs for rates and MOVER limits for the rate ratio.

**Table S5. Annual live-birth risk ratios for women with MS versus the matched comparison sample, by calendar period and adjustment model.**

| | Base | | | + Comorbidities | | | + SES | | | + Parity (Full) | | |
|---|---|---|---|---|---|---|---|---|---|---|---|---|
| | **RR** | **95% CI** | **p-value** | **RR** | **95% CI** | **p-value** | **RR** | **95% CI** | **p-value** | **RR** | **95% CI** | **p-value** |
| 2004-2008 | 0.76 | (0.70, 0.82) | <0.001 | 0.76 | (0.70, 0.83) | <0.001 | 0.78 | (0.72, 0.84) | <0.001 | 0.73 | (0.68, 0.79) | <0.001 |
| 2009-2013 | 0.86 | (0.81, 0.92) | <0.001 | 0.87 | (0.81, 0.92) | <0.001 | 0.87 | (0.82, 0.93) | <0.001 | 0.82 | (0.77, 0.87) | <0.001 |
| 2014-2018 | 0.81 | (0.76, 0.86) | <0.001 | 0.82 | (0.77, 0.87) | <0.001 | 0.83 | (0.78, 0.88) | <0.001 | 0.79 | (0.74, 0.84) | <0.001 |
| 2019-2022 | 0.75 | (0.70, 0.80) | <0.001 | 0.76 | (0.70, 0.81) | <0.001 | 0.77 | (0.71, 0.82) | <0.001 | 0.74 | (0.69, 0.80) | <0.001 |
| Interaction p-value | | 0.020 | | | 0.021 | | | 0.029 | | | 0.048 | |

Unit: woman-year; outcome: any live birth in the calendar year. Models are sequential: the base model adjusts for age group and region, with calendar period and an MS-by-period interaction in every model, and each later model adds the named covariates to the one before. SES: education and income. Robust standard errors clustered on woman. Interaction p: Wald test of equal risk ratios across the four periods.

**Table S6. Fully adjusted modified Poisson model for a live birth within a calendar year, all covariate estimates.**

| | Fully adjusted | | |
|---|---|---|---|
| | **RR** | **95% CI** | **p-value** |
| Calendar period, comparison sample | | | |
| 2004-2008 | | *Reference* | |
| 2009-2013 | 1.02 | (0.99, 1.05) | 0.18 |
| 2014-2018 | 0.99 | (0.96, 1.02) | 0.62 |
| 2019-2022 | 0.93 | (0.90, 0.95) | <0.001 |
| MS vs comparison, 2004-2008 | 0.73 | (0.68, 0.79) | <0.001 |
| MS vs comparison, 2009-2013 | 0.82 | (0.77, 0.87) | <0.001 |
| MS vs comparison, 2014-2018 | 0.79 | (0.74, 0.84) | <0.001 |
| MS vs comparison, 2019-2022 | 0.74 | (0.69, 0.80) | <0.001 |
| Age Group | | | |
| 18-19 | | *Reference* | |
| 20-24 | 5.00 | (4.15, 6.03) | <0.001 |
| 25-29 | 10.99 | (9.09, 13.28) | <0.001 |
| 30-34 | 11.47 | (9.49, 13.87) | <0.001 |
| 35-39 | 6.00 | (4.96, 7.27) | <0.001 |
| 40-45 | 0.96 | (0.79, 1.16) | 0.66 |
| Residence Region | | | |
| Stockholm | | *Reference* | |
| Östra Mellansverige | 0.99 | (0.96, 1.02) | 0.57 |
| Småland med öarna | 1.06 | (1.02, 1.10) | 0.005 |
| Sydsverige | 0.98 | (0.95, 1.02) | 0.32 |
| Västsverige | 1.00 | (0.97, 1.03) | 0.81 |
| Norra Mellansverige | 1.00 | (0.96, 1.04) | 0.96 |
| Mellersta Norrland | 1.06 | (1.00, 1.13) | 0.053 |
| Övre Norrland | 0.95 | (0.90, 1.00) | 0.040 |
| Depression | 0.87 | (0.82, 0.93) | <0.001 |
| Anxiety | 0.82 | (0.77, 0.87) | <0.001 |
| Diabetes | 0.82 | (0.72, 0.92) | 0.001 |
| Hypertension | 0.60 | (0.48, 0.75) | <0.001 |
| Obesity | 1.12 | (1.04, 1.21) | 0.003 |
| Thyroid disorder | 1.10 | (1.03, 1.17) | 0.005 |
| Epilepsy | 0.78 | (0.69, 0.89) | <0.001 |
| Highest education level | | | |
| <=9 years | | *Reference* | |
| 10-12 years | 0.94 | (0.90, 0.99) | 0.019 |
| >12 years | 1.15 | (1.10, 1.21) | <0.001 |
| Unknown | 0.53 | (0.41, 0.68) | <0.001 |
| Income quintile | | | |
| Q1 (lowest) | | *Reference* | |
| Q2 | 1.40 | (1.35, 1.44) | <0.001 |
| Q3 | 1.64 | (1.59, 1.70) | <0.001 |
| Q4 | 1.72 | (1.65, 1.78) | <0.001 |
| Q5 (highest) | 1.67 | (1.61, 1.74) | <0.001 |
| Parity (prior live-birth deliveries) | | | |
| Nulliparous | | *Reference* | |
| 1 | 1.71 | (1.66, 1.75) | <0.001 |
| 2 | 0.46 | (0.44, 0.47) | <0.001 |
| 3+ | 0.51 | (0.48, 0.54) | <0.001 |

Unit: woman-year; outcome: any live birth in the calendar year. Robust standard errors clustered on woman. Calendar-period rows: comparison sample relative to 2004-2008. Levels with fewer than 5 live births are fitted but not shown.

### RECORD checklist

**Manuscript:** Live Birth Rates Among Women With Multiple Sclerosis in Sweden, 2004 to 2022: A Population-Based Registry Study

Benchimol EI, et al. PLoS Med 2015;12(10):e1001885 (STROBE cohort items with RECORD extensions). All items are reported; exceptions are noted. Methods subsections: Data sources, Population-rate, Matched-cohort, Covariates, Statistical analysis. App, Supplementary Appendix; S, Supplementary Table.

| Item | Topic | Where reported |
|---|---|---|
| 1 | Design in title; informative abstract | Title; Abstract |
| 1.1 | Data type and databases named | Title; Abstract, Methods |
| 1.2 | Region and time frame | Title |
| 1.3 | Linkage stated | Abstract, Methods |
| 2 | Background and rationale | Introduction |
| 3 | Objectives | Abstract, Objective; Introduction, last sentence |
| 4 | Study design | Introduction, last sentence; Methods |
| 5 | Setting and dates | Methods, Data sources, Population-rate, Matched-cohort; App A1 |
| 6 | Eligibility, follow-up, matching | Methods, Data sources, Population-rate, Matched-cohort; App A1, A3 |
| 6.1 | Codes and algorithms for the population | Methods, Data sources; App A1 |
| 6.2 | Validation of codes | App A1 (no published validation; diagnostic-criteria field reported) |
| 6.3 | Linkage flow diagram | Not used; counts in Results, Cohort and deliveries |
| 7 | Variable definitions | Methods, Population-rate, Matched-cohort, Covariates, Statistical analysis; App A1, A4, A5; S1 |
| 7.1 | Codes for all variables | Methods, Data sources; S1; App A4, A5 |
| 8 | Data sources and measurement | Methods, Data sources, Covariates; App A2, A4, A5 |
| 9 | Bias | Methods, Matched-cohort, Statistical analysis; App A3; Discussion |
| 10 | Study size | Methods, Data sources; Results, Cohort and deliveries; App A3 |

| Item | Topic | Where reported |
|---|---|---|
| 11 | Quantitative variables | Methods, Covariates, Statistical analysis; Table 1; App A2, A4, A5 |
| 12 | Statistical methods, interactions, missing data, follow-up, sensitivity analyses | Methods, Covariates, Matched-cohort, Statistical analysis; App A3, A4, A6; S2 |
| 12.1 | Data access | Methods, Data sources; Data availability |
| 12.2 | Data cleaning | Methods, Covariates; App A1, A4, A5 |
| 12.3 | Linkage method and quality | Methods, Data sources; App A1, A3 |
| 13, 13.1 | Participants at each stage; selection | Results, Cohort and deliveries, Matched-cohort; App A3; S2 (no flow diagram) |
| 14 | Descriptive data, missing data, follow-up time | Tables 1 and 2; Results; App A4; S4 |
| 15 | Outcome data | Results; Table 3; Figure 1; S3, S4 |
| 16 | Main results, category boundaries, absolute measures | Results; Tables 1 to 3; Figure 1; S3 to S6 |
| 17 | Other analyses | Results; S5, S6; App A3, A6 |
| 18 | Key results | Discussion, first paragraph |
| 19, 19.1 | Limitations; implications of routinely collected data | Discussion |
| 20 | Interpretation | Discussion; App A6 (multiplicity) |
| 21 | Generalisability | Discussion |
| 22 | Funding | Funding statement |
| 22.1 | Protocol, data and code access | Data availability (no protocol registered) |
